# Perinatal suicide risk factors: A network analysis

**DOI:** 10.64898/2026.11.18.26368387

**Authors:** Kara Zivin, Chuwen Zhong, Lily Johns, Eskira Kahsay, Emma Spring, Anna Courant, Briana Mezuk

## Abstract

**Purpose:** Suicide during pregnancy or after birth (the perinatal period) represents a leading, preventable cause of United States maternal deaths. This study aimed to understand the complex relationship between perinatal status and life circumstances among perinatal suicide decedents in the United States and generate insights regarding suicide risk.

**Methods:** This cross-sectional study used decedent data from the 2003-2023 National Violent Death Reporting System suicide mortality registry (N=20,217). We applied psychometric network analysis using 18 binary life circumstances indicators representing four domains: relationships, health, socioeconomic status, and risk factors for suicide. We generated two networks for the overall sample and perinatal sample and assessed network structure, bridge centralities, and stability.

**Results:** The analytic sample included 1,297 perinatal and 18,920 non-perinatal decedents. The network of the overall sample was stable. Perinatal status did not appear at the center of the network but connected primarily with intimate partner problems. The network of the perinatal sample was modestly stable. Postpartum status was not at the center of the network but connected to depressed mood and mental health diagnoses. In both networks, depressed mood had the highest bridge betweenness, and there was a consistent link between physical health domains and socioeconomic domains.

**Conclusion:** Suicide prevention strategies targeting childbearing-aged women should incorporate factors beyond pregnancy, including physical health, intimate partner relationships, and socioeconomic challenges.

## INTRODUCTION

In the United States (U.S.), between 658 to 1205 women died of maternal causes each year between 2018–2022, with striking racial and ethnic inequities (1). The maternal mortality rate remains substantially higher in the U.S. than in similar countries (2). Importantly, maternal mortality review committees consider more than 80% of maternal deaths preventable, with perinatal suicide representing a leading, preventable cause of these deaths (3). Suicidal ideation (SI) is prevalent among perinatal individuals and recent population-level estimates point to a significant increase in the rates of perinatal SI in the last few decades (4–6). There is an urgent need for evidence-based strategies to prevent suicide during the perinatal period, particularly beyond traditional clinical factors such as psychiatric care (7).

Within a Life Course Framework, life transition events (e.g., marriage, job changes, etc.) are important for understanding suicide risk, as transition events may serve as stressors and increase the motivation for suicide (8). Pregnancy is one such event and the perinatal period, defined as pregnancy through one year after birth, is a distinct and vulnerable life stage. Specifically, women during the perinatal period are more likely to experience intimate partner problems, including intimate partner violence (4), a key interpersonal stressor often precipitating suicide (9)

Prior studies also show that a history of or current depressed mood, substance abuse, physical health problems, and recent bereavement (including but not limited to prior experiences of pregnancy loss) are associated with suicide during the perinatal period (10). More broadly, life stressors are understood as key risk factors for the onset or exacerbation of suicidal thoughts and behaviors (i.e., STBs), where such events intersect with existing psychological and interpersonal factors (e.g., history of depression, hopelessness) to confer added risk. Life stressors have also been identified as key determinants of perinatal mental health (11,12).

It is important to note that life circumstances seldom occur in isolation, especially in the perinatal period. For example, existing financial challenges can lead to unstable housing, which may in turn exacerbate intimate partner problems(8). Pregnant and postpartum women are uniquely vulnerable to financial and relationship stressors as they may experience pregnancy-related health challenges and may experience greater employment instability (12). This cascading pattern reflects the Life Course Framework’s view that stressors accumulate and interact over time rather than acting independently.

Therefore, preventing suicide during the perinatal period requires a more nuanced understanding of pregnancy within the broader context of overall life circumstances. However, there is a lack of empirical evidence on how life circumstances from different domains relate to one another. Most existing studies of perinatal suicide risk focus on a specific exposure, such as intimate partner violence, or rely on an aggregated index (e.g., the number of circumstances a case reported) (13). These approaches make it difficult to represent and describe how individual life circumstances interrelate. Understanding the structure of the interrelated relationships between individual life circumstances from different domains can situate pregnancy within the social, relationship, and economic context, which can inform intervention beyond traditional clinical factors.

Network analysis offers a promising alternative analytic approach that can quantify and visualize complex, multidimensional relationships among multiple life circumstances simultaneously (14). Because such stressors accumulate and interact over time, a network approach is well suited to mapping how circumstances connect and cluster to shape risk during the perinatal transition. It has been applied widely in clinical psychology to understand interrelationships between symptoms of mental health conditions (15,16) and the between life stressors (e.g., childhood trauma) and psychosis symptoms (17). Applying this network approach to perinatal suicide can generate new insight into how circumstances cluster and connect among women who die by suicide during this period. Using network analysis, this study aimed to visualize and explore relationships among life circumstances for female suicide decedents of childbearing age (10-50 years old) and, separately, for female suicide decedents who died during the perinatal period. Specifically, we examined 1) whether circumstances from different domains are connected and how, 2) whether perinatal status is a key circumstance and which circumstances it links to, and 3) which circumstances are most central in the network. With the lens of network analysis, findings may offer new insights to inform research design and interventions aimed at preventing perinatal suicide deaths.

## METHODS

### Data source

This study analyzed suicides and deaths of undetermined intent using data from the National Violent Death Reporting System (NVDRS) from January 1, 2003 to December 31, 2023. The NVDRS is an active, state-based surveillance system managed by the CDC that collects data on violent deaths occurring in the United States and is an important resource for suicide prevention research (18,19). Since 2019, NVDRS includes data from all 50 states, the District of Columbia, and Puerto Rico, though coverage years vary by state (19). Notably, the NVDRS Restricted Access Database contains text narratives (150-300 characters) and life circumstance variables collected from coroner/medical examiner and law enforcement reports, death certificates, suicide notes, and interviews with decedents’ family and friends. We included both suicides and deaths of undetermined intent to capture potential misclassification (20,21). The NVDRS used International Classification of Diseases, Tenth Revision, Clinical Modification codes to identify suicide (X60-X84, Y87.0, and U03) and undetermined death (Y10-Y34, Y87.2, and Y89.9).

Due to the use of decedent data, the University of Michigan Institutional Review Board exempted this study from review and waived the need for informed consent. We followed the Strengthening and Reporting of Observational Studies in Epidemiology (STROBE) reporting guidelines (22).

### Sample

The analytic sample included female decedents of childbearing age (10-50 years old) with complete data for demographics, pregnancy status at time of death (i.e., currently pregnant, 0-42 days post-partum, 43 days to 1 year postpartum), and known life circumstances. Figure 1 provides a flowchart of sample selection.

**Figure 1.**
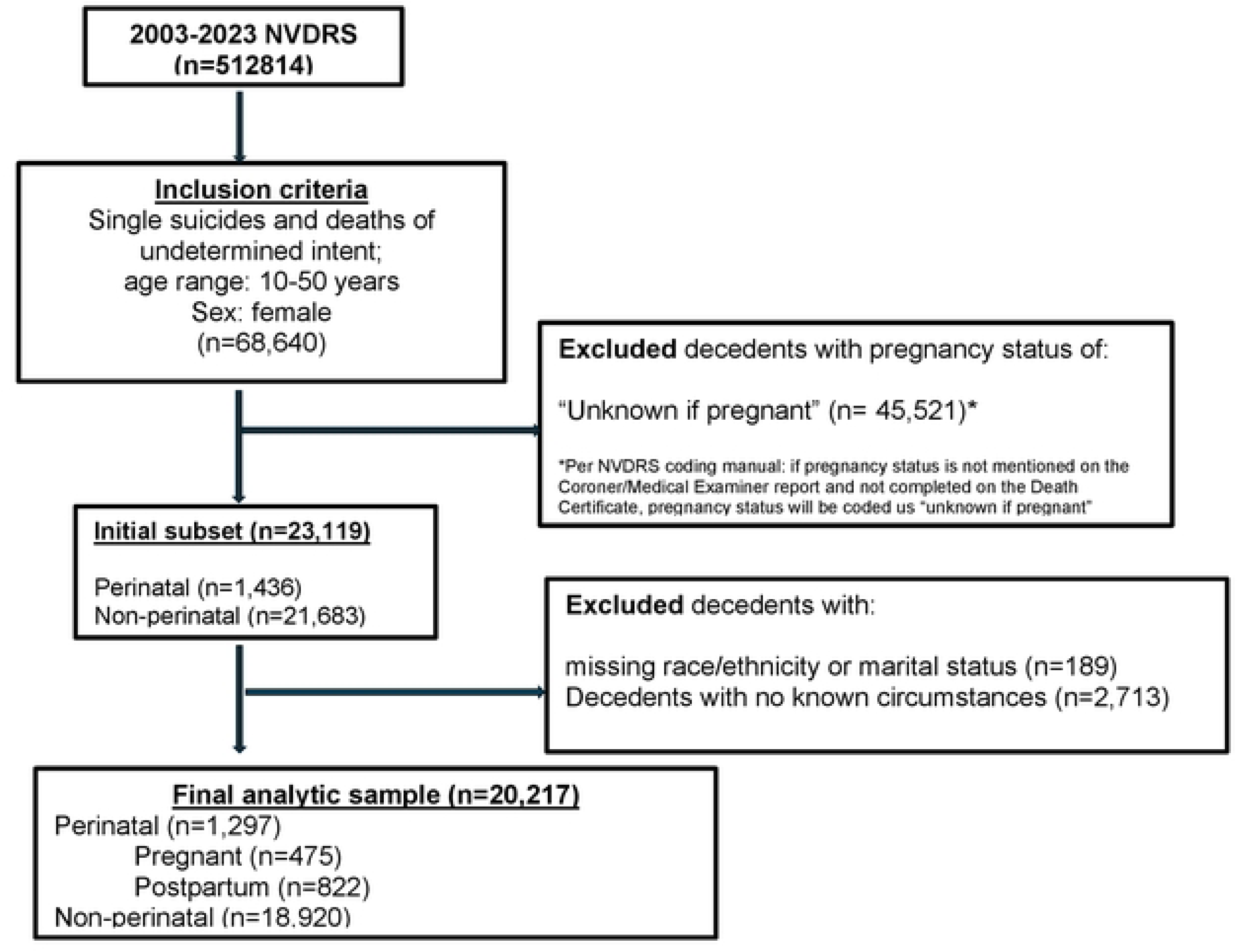
Flowchart of analytic sample selection, National Violent Death Reporting System (2003-2023). Figure 1 provides the flowchart of the analytic sample selection. We excluded decedents with unknown pregnancy status, and decedents with missing race/ethnicity or marital status or known circumstances.

### Conceptualization of Pregnancy Status

The original pregnancy status variable has four categories: “not pregnant within last year”, “pregnant at the time of death”, “Not pregnant but pregnant within 42 days of death”, and “Not pregnant but pregnant within 43 days to 1year of death”. We derived two binary variables from the original coding. The first binary variable is “perinatal status” where: decedents who had “not pregnant within the last year” were coded as “non-perinatal”, and the remaining decedents were coded as “perinatal”. We used the “perinatal status” to generate two nested samples: (1) all female decedents of childbearing age, including both perinatal and non-perinatal decedents (referred to hereafter as the overall sample) and (2) all perinatal decedents. We also included “perinatal status” as a node in the network for the overall sample. Among the perinatal sample, we derived a second binary variable, “postpartum status”, where: decedents who were “pregnant at time of death” were coded as “non-postpartum” and the remaining decedents were coded as “postpartum”. We included “postpartum status” as a node in the network for the perinatal sample. Appendix 1 describes full operationalization.

### Circumstances variables

NVDRS includes life circumstances as binary variables (“yes” vs. “no, not available, or unknown”) and codes as “yes” if the circumstance was perceived as contributing to the death. This rule does not apply to “mental health diagnosis” and “substance abuse;” NVDRS codes these two variables as “yes” regardless of direct contribution to the death (23). Informed by the life-course framework of suicide (8), we identified 18 salient circumstances variables from four domains: interpersonal relationships (intimate partner, family relationship, other relationship, argument, and history of abuse); health (perinatal status/pregnancy status, mental health problem, recent treatment non-adherence, and physical health problem); socioeconomic status (job problem, financial problem, and eviction from house); and established risk factors (depressed mood, history of suicide attempt, history of suicide thoughts, any addiction, recent family or friend suicide, and other family or friend death). We selected circumstances variables collected since 2013 and discard those collected in more recent years to maximize data availability. For circumstances that showed higher correlation than others (such as other family or friend death and traumatic anniversary), we kept the circumstance with higher counts. Appendix 2 details all circumstances, and Appendix 3 presents a correlation heatmap for pairs of circumstances.

### Network construction

We used psychometric network analysis to visualize and characterize relationships among life circumstances. In general, a network consists of two elements: “nodes” (the points in the network) and “edges” (the lines between points in the network) to represent the associations between a collection of features (e.g., individuals, risk factors, symptoms) (14).

In this study, nodes represent 18 individual circumstances, color-coded to indicate four domains. The edges of the network represent the association between pairs of circumstance variables, and the color and width of edges indicate the orientation and strength of the corresponding association, respectively (16).

Since all nodes were dichotomous, we employed the Ising Model and the widely used regularization approach the Least Absolute Shrinkage Operator (LASSO) with Extended Bayesian Information Criterion (EBIC) to estimate edge weights (24–26). This approach ensures that the most important edges remain while weights of weak edges shrink to zero and drop from the network, resulting in a more interpretable network. The EBIC hyperparameter γ controls network density by determining the number of edges to shrink. Therefore, the higher the γ, the more the edges shrink, and the sparser the remaining network (26). We generated two networks, one for the overall sample and one for the perinatal sample. We set γ to 0.25 for the network of the overall sample, as recommended in prior literature (26). Due to the sample size of the perinatal subgroup being only 1/16 of the overall sample, a network with γ to 0.25 was less informative. We reports the network with γ to 0.1 for the perinatal subgroup and provide the network with γ to 0.25 in the Appendix 4. Both networks did not adjust for demographic covariates to align with the analysis purpose of understanding the network structure and relative interaction between nodes in the respective samples.

### Statistical analysis

We conducted descriptive analyses and used Student t-tests (for continuous variables) and Chi-square tests (for categorical variables) to compare demographic characteristics between perinatal and non-perinatal decedents and between postpartum and currently pregnant perinatal decedents.

To evaluate the overall connectivity of the network, we calculated the network density. The network density is the number of observed edges divided by all possible edges, ranging from 0 to 1, in where 0 indicates that all nodes appear isolated from all others and 1 indicates that all nodes connect to all others. We used bridge centrality metrics (i.e., bridge strength and bridge betweenness) to quantify how well nodes of one domain (e.g., socioeconomic status) connect to nodes from other domains (e.g., health); through these metrics, we aimed to identify key nodes that bridged different network domains (27). Specifically, bridge strength represents the sum of absolute weights of all edges between one node and nodes from different domains, capturing direct cross-domain connection of that node. Bridge betweenness represents the number of times that one node occurs on the shortest path between nodes from two different constructs, reflecting the indirect gatekeeper role of a node. We assessed network stability using two approaches (25). First, we performed non-parametric bootstrap (n=2,000) to obtain bootstrapped confidence intervals (BCI) to assess the reliability of the edge estimates. Second, we performed a case-dropping bootstrap (n=2,000) to assess the stability of bridge centrality metrics with the Correlation Stability (CS) Coefficient. Appendices 5 and 6 describe these methods in detail.

### Sensitivity analysis

In the preliminary investigation, we found that the 18-node network for the perinatal sample appeared highly unstable. As a result, we could not interpret that network because edges and patterns could have arisen due to change. Small sample size and low variance of some nodes may drive this high instability. A node has low variance if most of the observations have the same value (in our case, “no, not available”). Nine out of 18 nodes had only 2% (29 cases) to 5% (110 cases) of the “yes” count. To improve the network stability and interpretability, we generated a network with 13 nodes by 1) combining related circumstances (“job problem” and “financial problem” to “job/financial problem”; “friend/family suicide” and “other death of friend/family” to “other deaths”) and 2) removing two nodes (“treatment non-adherence” and “ history of child abuse”) that had a low “yes” count and did not connect to other nodes in the early investigation. We reported on the 13-node network for the perinatal sample and provided information for the 18-node network in the Appendix 7.

Both the overall and perinatal sample networks had an edge between the circumstances of physical health problems and job/financial problems. Given that physical health problems remain relatively rare for women of childbearing age compared to mid-life and older women, we did not expect this finding (28). Therefore, three reviewers (authors L.J., E.S., and C.Z) conducted a post-hoc review of a random sample of NVDRS text narratives with both physical and job/financial problems (overall sample N=25 and perinatal sample N=22). As detailed elsewhere, NVDRS abstractors write text narratives to provide additional details about circumstances in the decedent’s life at the time of death not fully captured by binary circumstance variables (29). This post-hoc analysis thus provides additional contextual information about the nature of the observed linkage between physical health and job/financial problems identified in the networks.

We used RStudio (Version 2024.12.1+563) for data management and network analysis. Appendix 9 details the packages used. We performed analyses between November 2024 and February 2026.

## RESULTS

### Descriptive analysis

Between 2003 and 2023, there were 1,297 perinatal and 18,920 non-perinatal suicide and undetermined intent decedents in NVDRS. Compared to non-perinatal decedents, perinatal decedents were significantly younger (29.2 years old vs. 35.6 years old), less likely to be White (66.4% vs. 79.2%), less likely to have some college or higher educational attainment (38.0% vs. 43.1%), and more likely to be either married (33.5% vs 27.6%) or single/never married (51.5% vs 43.8%). Among perinatal decedents, 822 were postpartum, and 475 were currently pregnant at the time of death. Postpartum and pregnant decedents were similar in age, race/ethnicity, and educational attainment. Postpartum decedents were slightly more likely to be married (36.1% vs 29.1%). Additional characteristics appear in Table 1.

**Table 1.** Demographic characteristics of female decedents by pregnancy status within one year using NVDRS data from 2003-2023 and statistical tests.

|  | **Decedents in childbearing age** |  |  | **Postpartum and pregnancy decedents** |  |  |
|---|---|---|---|---|---|---|
|  | **Non-perinatal** | **Perinatal** | **Test Statistics (p-value)** | **Postpartum** | **Pregnant** | **Test Statistics (p-value)** |
|  | **(n=18,920)** | **(n=1,297)** |  | **(n=822)** | **(n=475)** |  |
| **Age, Mean(SD)** | 35.6 (10.8) | 29.2 (7.3) | 29.7 (<.0001) | 29.00 (7.1) | 29.56 (7.6) | -1.3(0.19) |
| **Race/Ethnicity, n(%)** |  |  | 120.0 (<.0001) |  |  | 3.7(0.30) |
| Hispanic -1 | 1,485 (7.8) | 158 (12.2) |  | 103 (12.5) | 55 (11.6) |  |
| Non-Hispanic Black -2 | 1,343 (7.1) | 159 (12.3) |  | 90 (10.9) | 69 (14.5) |  |
| Non-Hispanic White-3 | 14,993 (79.2) | 861 (66.4) |  | 552 (67.2) | 309 (65.1) |  |
| Others-4 | 1,099 (5.8) | 119 (9.2) |  | 77 (9.4) | 42 (8.8) |  |
| **Educational Attainment, n(%)** |  |  | 48.5 (<.0001) |  |  | 4.7(0.20) |
| <= 12th grade 1 | 3,401 (18.0) | 186 (14.3) |  | 116 (14.1) | 70 (14.7) |  |
| HS/GED2 | 5,884 (31.1) | 465 (35.9) |  | 288 (35.0) | 177 (37.3) |  |
| Some college/college and above3 | 8,151 (43.1) | 493 (38.0) |  | 329 (40.0) | 164 (34.5) |  |
| Unknown 4 | 1,484 (7.8) | 153 (11.8) |  | 89 (10.8) | 64 (13.5) |  |
| **Marital status, n(%)** |  |  | 112.9 (<.0001) |  |  | 7.1(0.03) |
| Married | 5,225 (27.6) | 435 (33.5) |  | 297 (36.1) | 138 (29.1) |  |
| Single/Never married | 8,286 (43.8) | 668 (51.5) |  | 410 (49.9) | 258 (54.3) |  |
| Divorced/Separated/Widowed | 5,409 (28.6) | 194 (15.0) |  | 115 (14.0) | 79 (16.6) |  |
| **Other covariates** |  |  |  |  |  |  |
| Injured at Victim's Home (Yes) | 1,5074 (79.7) | 1011 (77.9) | 2.1 (0.15) | 641 (78.0) | 370 (77.9) | 0.00 (1.00) |
| Autopsied (full or partial) | 12,183 (64.4) | 862 (66.5) | 2.2(0.14) | 516 (62.8) | 346 (72.8) | 13.2 (0.003) |
| In Labor force (Yes) | 6,717 (35.5) | 486 (37.5) | 2.0(0.16) | 328 (39.9) | 158 (33.3) | 5.4(0.02) |

### Network for the overall sample

Figure 2a shows the overall sample network. The network density was 0.46 (70 observed edges/153 potential edges), indicating a fairly high level of interconnection of life circumstances across life domains. The BCIs appeared reasonably narrow and did not cross zero, indicating stable results (Appendix 5a). Both bridge centrality metrics were stable, and all Correlation Stability (CS) coefficients were 0.75, indicating that we could drop at least 75% of data to maintain a correlation of 0.7 with 95% certainty with the observed dataset (Appendix 5b). This suggests that both bridge centrality metrics are robust.

**Figure 2.**
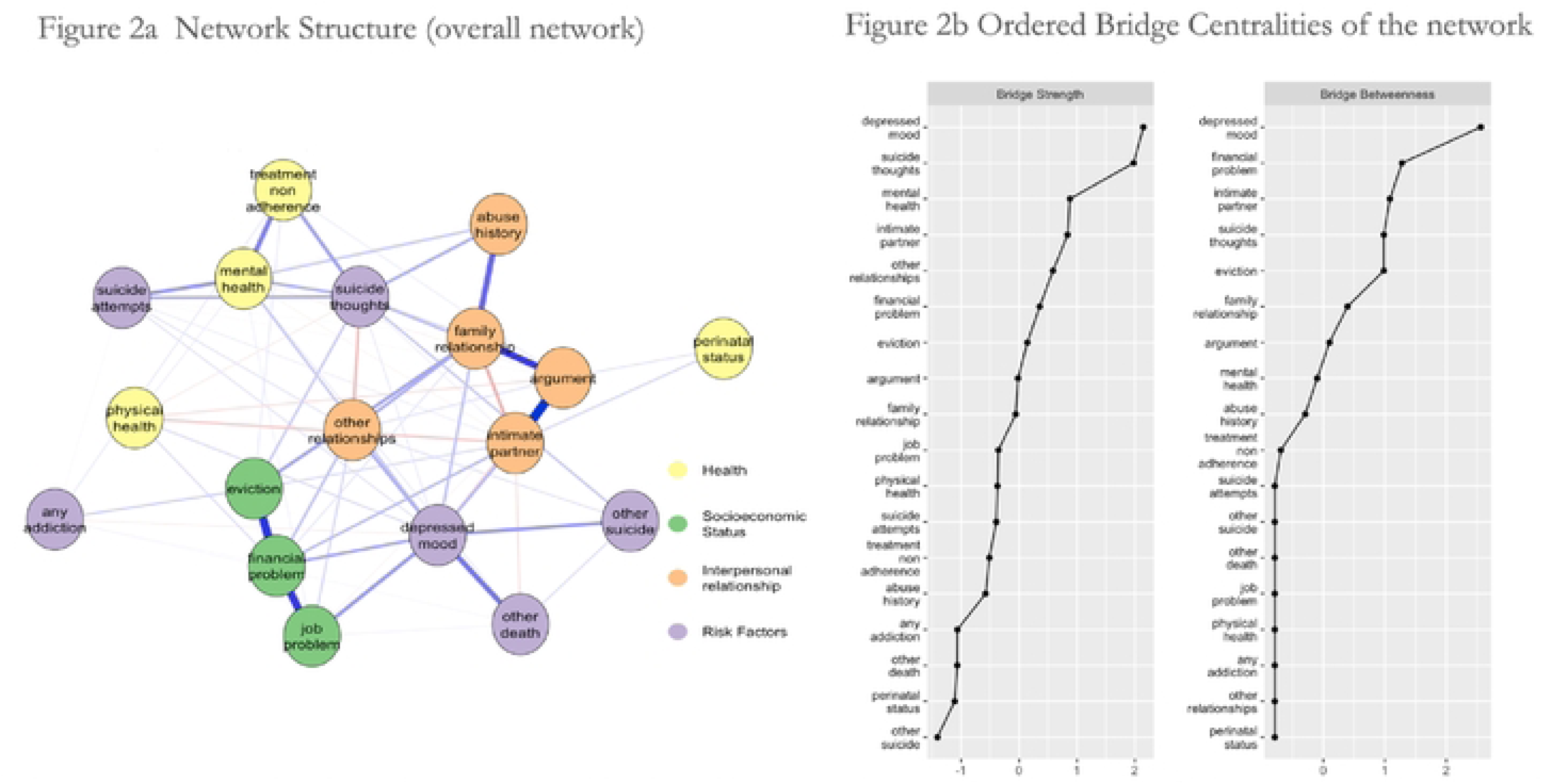
Network of the circumstances and ordered bridge centrality (18 nodes) of the overall sample, National Violent Death Reporting System (2003-2023) (n=20,217) In Figure 2a, colors indicate domains of nodes: light yellow: Health; light green: Socioeconomic Status; orange: Interpersonal relationship; purple: Risk factors. Positive edges are blue, negative edges are red. The darker the edge, the stronger the connection. In Figure 2b, nodes are ordered based on their bridge centrality values. Bridge strength is the sum of absolute weights of all edges between a node and nodes from different domains; bridge betweenness is the number of times a node is on the shortest path between nodes from two different constructs.

The network illustrates five features of interrelationships between these life circumstances. First, perinatal status appeared at the periphery of the network, with only two positive edges linking perinatal status to the network through the nodes intimate partner problem and argument. Second, nodes from the interpersonal domain (e.g., argument, family relationship problem, and intimate partner problem) occupied central positions in the network. Third, the health domain connected to the socioeconomic domain; specifically, physical health problems had an edge with financial problems. Fourth, unexpectedly, mental health problems did not directly connect to addiction problems. Finally, Figure 2b displays bridge centrality indices for this network, illustrating that risk factor domain nodes, depressed mood and SI, and the socioeconomic domain node financial problem had the highest bridge centrality values and therefore represent key nodes that bridge across domains.

### Network for the perinatal subgroup

Figure 3a presents the 13-node perinatal network. As expected, given the smaller sample size, the network appeared sparser than the overall network, with a network density of 0.28 (22 observed edges/78 potential edges). The BCIs appeared reasonably narrow and did not cross zero, indicating edges were modestly stable (Appendix 6a). The CS coefficients for bridge betweenness and bridge strength were 0.36 and 0.05 (<0.25 cutoff), respectively, indicating that bridge betweenness was modestly stable, whereas bridge strength was unstable and not interpretable (Appendix 6b). The 13-node perinatal network had structure similar to the 18-node perinatal network (Appendix 7).

**Figure 3.**
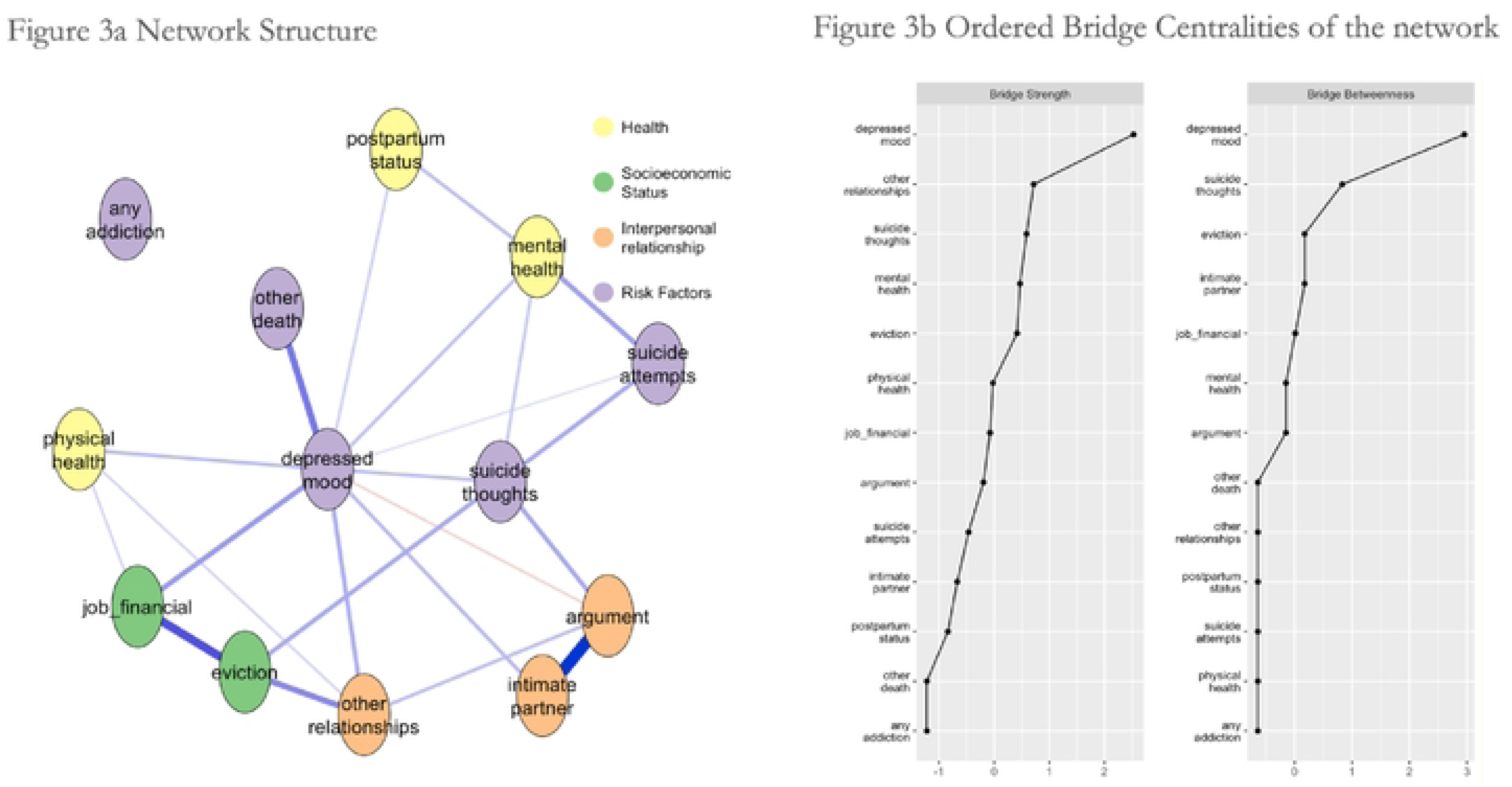
Network of the circumstances and ordered bridge centrality of the perinatal sample (13 nodes), National Violent Death Reporting System (2003-2023) (n=1,297) In Figure 3a, colors indicate domains: light yellow: Health; light green: Socioeconomic Status; orange: Interpersonal relationship; purple: Risk factors. Positive edges are blue, negative edges are red. The darker the edge, the stronger the connection. In Figure 3b, nodes are ordered based on their bridge centrality values. Bridge strength is the sum of absolute weights of all edges between a node and nodes from different domains; bridge betweenness is the number of times a node is on the shortest path between nodes from two different constructs.

We observed the following key findings. First, postpartum status appeared peripherally in the network. Second, postpartum status had two edges linking to mental health and depressed mood. Third, risk factor domain nodes, such as depressed mood, suicide thoughts, appeared centrally. Fourth, the physical health node linked with job/financial problems. Finally, depressed mood and suicide thoughts had the highest bridge betweenness, representing key bridges between different domains, as presented in Figure 3b.

### Post-hoc analysis of narrative texts

In the overall and perinatal sample networks, the linkage between physical health problems and socioeconomic domain nodes was likely bidirectional. In some narratives, ongoing physical health problems exacerbated existing life stressors such as intimate partner problems and substance use, which led to job loss and subsequent financial hardships. In other narratives, financial strain intensified existing physical health problems. In the overall sample, chronic illness and family-role obligations occurred centrally in the overall narratives, whereas in the perinatal sample, chronic pain and pregnancy-related financial stressors appeared most salient and frequently interacted with intimate partner violence. Appendix 8 presents abbreviated and edited illustrative narratives.

## DISCUSSION

This study used network analysis to visualize and explore the inter-relationships between multiple domains of life circumstances among female suicide decedents, with an emphasis on the perinatal period. Guided by the understanding that pregnancy is not an isolated event but rather occurs within the context of prior and concurrent life experiences, we aimed to understand how perinatal status or postpartum status connect to other life circumstances, how circumstances from different life domains relate to one another, and which circumstances serve as key nodes linking different life domains among a large national sample of female suicide decedents of childbearing age.

In the overall sample network, we noticed that perinatal status did not appear as a central node. This finding reflects that perinatal status did not represent a key connector to other life circumstances. Perinatal status connected to the overall sample network through limited linkages via interpersonal relationship factors (i.e., intimate partner problems and arguments). Although interpersonal problems did not appear in highly central positions in the perinatal sample network, they directly connected to depressed mood and SI nodes, which represented the most influential nodes in the network.

Prior research offers support for this linkage between intimate partner problems and perinatal suicide. The interpersonal theory of suicide posits that a reduced sense of belonging, facilitated by factors such as interpersonal conflict, contributes to suicide risk (30). During the perinatal period, self-identity and relationships evolve (31), which can lead to heightened salience of interpersonal relationship factors. Prior studies identified a dose-response relationship between social conflict and increased prenatal depression, with more interpersonal risk factors contributing to greater depression risk (32). Further, the Life Course Framework, applied to the perinatal period, illustrates that maternal suicide risk can peak when the stressors of new parenthood intersect with a substantial decline in screening and support in both medical and social contexts. This framework indicates that vulnerability fluctuates as early protective monitoring and support gives way to subsequent isolation and role strain during the first postpartum year. As such, this period represents significant changes in both motivation for suicide and more drastic changes in opportunity of attempting suicide relative to other periods of the life-course (8,33). In addition, interventions focused on building interpersonal skills, decreasing interpersonal conflict, and increasing interpersonal support significantly reduced prenatal depression rate compared to usual care (OR 4.99; 95% CI: 2.08-11.97) (34). Our findings offer further support for the association between depression and interpersonal risk factors and suicide during for women during the perinatal period.

Depressed mood and SI had the highest bridge centrality both in the overall sample and perinatal sample networks, indicating that these risk factors represent important leverage points for clinical intervention. Beyond clinical benefits of detection and timely and effective treatment of depressed mood and suicidal thoughts, this analysis suggests that addressing these risk factors may yield broader benefits due to their linkages to other domains. For example, in the perinatal network, depressive symptoms associated with job/financial strain, intimate partner problem, physical health conditions, and postpartum status (35,36).

Although the present study was cross-sectional and did not identify causal relationships, our findings of interconnectedness of life circumstances align with interventional studies of perinatal outcomes evaluating comprehensive approaches to addressing key suicide risk factors such as depression both during and after the perinatal period. For example, the collaborative care model, which involves providing key interpersonal support (e.g., nurse care manager or community health worker) improves depression outcomes across a wide array of patient populations, including pregnant women, relative to usual care (37). Similarly, group prenatal care models are associated with improved psychosocial outcomes, in part due to their effectiveness at fostering community, increasing social support, and reducing isolation (38). These findings also emphasize the importance of access to programs such as the Special Supplemental Nutrition Program for Women, Infants, and Children (WIC) that relieve financial stress and improve maternal mental health (39,40). Other recent research supports this point, suggesting that newer access programs that offer community-wide, unconditional prenatal and infant cash transfers also improve economic stability, mental health, and well-being (41).

Finally, the links between physical health problems and financial problems, while well-established in other populations (42), remain underexplored as contributors to suicide among adolescent and young adult women. Future research should focus on more clearly understanding this relationship within the perinatal population.

### Limitations

Data abstractors enter NVDRS data from original death investigation records. Therefore, both under-documentation and variability in reporting across states, which differ in their death investigation systems, can occur. We attempted to minimize variability by limiting our node set to circumstances consistently recorded in this registry since 2013. However, to maximize sample size (particularly for the perinatal sample), our data spanned 20 years (from 2003 to 2023). Importantly, absent circumstances do not definitively indicate that those circumstances did not occur, simply that they were not reported. Therefore, biases in the assessment of circumstances and data availability limit the generalizability of findings. Moreover, the relatively small proportion of decedents classified as perinatal (6% of the overall sample) may have contributed to an imbalanced distribution of this node and influenced its position within the overall network. Lastly, the cross-sectional nature of this analysis precludes causal inference. The results describe associations between circumstances and do not imply causal pathways.

## CONCLUSION

Every pregnancy occurs within the prior context of a life and concurrent interpersonal relationships and socioeconomic challenges. This network analysis of perinatal suicide risk factors revealed intersections between perinatal suicide and factors beyond the pregnancy experience itself, including intimate partner problems, physical health conditions, and financial challenges. Efforts to improve perinatal mental health and prevent perinatal suicide deaths will benefit from collaboration from policymakers, healthcare systems, and community-based organizations to address the interconnected nature of mental health, interpersonal problems, and socioeconomic strain.

## Disclosures of Potential and Actual Conflicts of Interest

The authors have no conflicts of interest to disclose.

## Data Availability

No data were generated by this study. Data used in this analysis are available upon request from the CDC through their restricted-access data process. Our use of these restricted-access NVDRS data is governed by a Data Use Agreement (DUA) with the CDC. This DUA legally prohibits us from sharing these data with outside investigators. Other investigators can contact to gain access to these restricted-access NVDRS data. Investigators should follow the procedures outlined here: https://www.cdc.gov/violenceprevention/datasources/nvdrs/dataaccess.html. Other NVDRS data are publicly available: https://www.cdc.gov/violenceprevention/datasources/nvdrs/datapublications.html.

https://www.cdc.gov/violenceprevention/datasources/nvdrs/datapublications.html

